# On Estimating Age and Gender from Parkinson’s Disease Diagnostic-Oriented Recordings Using Wav2Vec 2.0

**DOI:** 10.64898/2025.12.29.25343161

**Authors:** Ondrej Klempir, Ales Tichopad, Radim Krupicka

## Abstract

Self-supervised speech foundation models (SFMs) are increasingly applied in biomedical research, yet it remains unclear to what extent demographic attributes such as age and gender are encoded in pathological speech. This study evaluates a pretrained Wav2Vec 2.0 SFM with two custom prediction heads for age and gender estimation, without exposing the model to any data from the evaluated cohorts. Experiments were conducted on three independent multilingual datasets comprising 244 subjects, including healthy controls, Parkinson’s disease (PD) patients, and related parkinsonian syndromes. Multiple speech tasks, read text, diadochokinesis, and sustained vowel phonation, were evaluated using gender classification accuracy, correlations with chronological age, and group-level statistical analyses. Compared to a Wav2Vec 2.0 XLSR feature-extraction baseline, the primary presented approach achieved consistent performance improvements of at least 8%. Gender estimation was highly robust, reaching 94–100% accuracy across datasets, tasks, and diagnostic groups. Age estimation showed significant correlations with true age for connected speech, including PD speakers, but failed for sustained vowel phonation across all groups, revealing systematic age bias. These findings demonstrate that pretrained SFMs robustly encode gender and preserve age-related structure in connected pathological speech, while exposing task-dependent limitations for age estimation. The results support the use of SFMs for demographic metadata extraction in clinical speech analysis and highlight the need to monitor age-related bias in diagnostic-oriented speech tasks.

## 1. Introduction

Self-supervised foundation models have been rapidly reshaping how healthcare can leverage unstructured data. In speech research, these models are capable of learning powerful acoustic and prosodic representations directly from large and heterogeneous audio collections, enabling downstream systems to detect clinical signals that previously required handcrafted features or task-specific modeling. Speech foundation models (SFMs), such as Wav2Vec 2.0 or HuBERT, have shown strong performance in Parkinson’s disease (PD) and other voice disorders detection and can outperform traditional acoustic feature sets (Cai et al., 2024; Favaro et al., 2023). PD is the second most common neurodegenerative disorder after Alzheimer’s disease (AD). It primarily affects individuals aged 65 years and older (Morris, 2000). PD cases are expected to double by 2030 (since 2005) and continue a sharp growth trajectory into the following decade (Bhidayasiri et al., 2023).

The recent discussion and corresponding study on the usefulness of speech data for developing classifiers for PD detection (Zhong et al., 2025), demonstrated the benefits of balancing participants’ gender and disease-status distributions. However, the corpora obtained either from a publicly or internally available source may lack reliable demographic metadata. This might pose a problem, as age and gender can affect baseline acoustic characteristics and may interact with disease-related changes. When such information is missing, models may theoretically exploit correlations with demographics instead of pathology, especially given that PD cohorts are typically older while healthy controls (HC) are often younger. Without addressing these imbalances, reported accuracy may reflect demographic confounds rather than true clinical discriminability. These concerns are amplified by the behavior of recent foundation models. Being trained on large, heterogeneous speech corpora, they naturally cover demographic structure. When applied to imbalanced datasets as feature extractors, downstream classifiers may rely on age or gender cues rather than PD-specific voice patterns. This shortcut learning can artificially inflate performance, particularly in designed binary contrasting (HC vs. PD). Therefore, in case when demographic variables are not available, at least coarse estimation or characterization might be of interest to identify potential confounders, support external validation, and enable transparent reporting.

Speech-based PD detection can be influenced by speaker gender, age, language, recording device, task characteristics and other factors. Evidence showed that sex (biological attributes) affects disease-related voice changes, with males often exhibiting greater impairments (Cao et al., 2025). Recording conditions and the choice of diagnostic-oriented tasks can substantially alter the acoustic footprint and downstream performance. To address this partially, several robust, language-independent acoustic features relevant to PD detection have already been identified (Scimeca et al., 2023). However, all of these factors (and not limited to) underscore the need to understand how they possibly interact with foundation model behavior, with age and gender standing out for considering possible demographics differences.

### 1.1. Research objectives

#### What is already known on this topic and What this communication adds

While gender and age prediction works reliably in healthy speech, it remains unclear whether this capability extends to various types of pathological recordings. Clinical assessment of speech disorders typically involves standardized tasks, yet many recordings or entire datasets lack demographic annotation. Our aim is therefore to assess whether a single, ready-to-use foundation model can provide consistent demographic estimates across heterogeneous PD-related speech material, supporting clearer dataset characterization and more reliable downstream model evaluation.

The overarching objective of this research is to provide data-driven evidence addressing the following central question: “Can self-supervised SFMs be used for automatic extraction of patient metadata, even in the absence of prior demographic information and when speech is affected by pathology?”.

Despite the growing adoption of SFMs in biomedical speech and voice analysis, current research predominantly focuses on binary prediction tasks, such as disease detection. As a result, there remains a limited understanding of:

- how these models operate beyond disease classification,
- how adaptable they are to a broader range of downstream tasks, such as demographic or metadata estimation,
- and what intrinsic information they encode when pathological speech is not explicitly represented in their pretraining data.

Furthermore, it is still unclear under what conditions SFMs possibly underperform, and what capabilities emerge from their learned global representations, particularly in realistic clinical scenarios where curated pathological recordings are scarce or unavailable during pretraining. To address these gaps, this research aims to systematically investigate the behavior and limitations of SFMs across diverse diagnostic-oriented pathological speech recordings.

### 1.2. Research contributions

The key contributions of this work can be summarized as follows:

#### 1. Comprehensive evaluation of SFM for patient metadata estimation

Through extensive experiments, we demonstrate that the self-supervised foundation model Wav2Vec 2.0 can effectively estimate age and gender from connected speech. We report the results across three multilingual corpora, including HC, PD patients, and related syndromes.

#### 2. Baseline pipeline for comparison

To contextualize the presented approach, we introduce a baseline method based on an established pipeline that employs pretrained speech models solely as fixed feature extractors, followed by conventional downstream modeling. The primary approach being investigated consistently achieved substantial improvements over this baseline.

#### 3. Theoretical insight into task-dependent limitations of SFM

We provide evidence that SFM exhibits limited capability to accurately model certain tasks, particularly predicting age based on sustained vowel phonation.

## 2. Related work

Foundation models can generally predict speaker gender and age with high accuracy in healthy speech (Burkhardt et al., 2023; Truong et al., 2022; Yang et al., 2025), which is expected for clean, normative recordings. At least a few pre-trained, openly available models hosted on HuggingFace, e.g. (Burkhardt et al., 2023; Common-Voice-Gender-Detection at Hugging Face, 2020; Desplanques et al., 2020), report near-perfect gender-prediction accuracy and also possibly support age estimation. These models require no additional training and are immediately usable, making them candidates for demographic characterization. However, for disordered speech evidence remains limited. Although many PD assessment protocols use standardized tasks (e.g. fixed reading passages, monologues, diadochokinetic tasks, sustained vowels), little is known about how well foundation models infer demographic attributes across these task types when pathology is present. Our goal is to evaluate whether such “ready-to-go,” multipurpose models remain reliable when applied to PD-related speech.

In the prior work (Klempíř & Krupička, 2024), Wav2Vec 1.0 was used to predict age in an Italian dataset that included PD participants during a standardized reading task. The model captured age structure with reasonable accuracy, effectively separating younger from older speakers despite pathological variation. While gender can often be inferred perceptually, foundation models offer an opportunity to objectivize this process, reduce listener bias, and support reproducible demographic characterization. To provide an example, recent studies have reported the gender distribution of the Mobile Device Voice Recordings at King’s College London (MDVR-KCL) English corpus (Jones et al., 2025; Kim et al., 2024), despite the dataset lacking official demographic metadata. To our knowledge, these studies did not describe how gender was inferred, and it is anticipated that classifications were made by listening. These assumptions highlight the need for systematic, model-based demographic estimation in PD speech datasets.

Existing relevant research found that demographic cues remain detectable even in pathological speech. The study (Gómez-García et al., 2015), demonstrated that age classification (adult vs. older adult) is feasible using sustained vowels, showing that age structure can be recovered even in disordered voices and suggesting a path toward age-aware pathology models. For gender, the “Octopus” vowel-based PD and gender recognition study (Tuncer & Dogan, 2019) reported an average accuracy of 93.8% across six classifiers using a vowel dataset (three vowels per person, 252 speakers). More recently, GeHirNet (Wu et al., 2025) proposed a two-stage, gender-aware pathology classifier trained on multiple pathological speech corpora, including the gender-balanced PC-GITA PD dataset. Although the method achieved high accuracy (97.6%) by modeling gender cues in Stage 1 and pathology in Stage 2, it requires end-to-end training, focuses on pathology detection rather than standalone demographic estimation, and is limited to sustained vowel /a/. To our knowledge, no study directly and comprehensively evaluated the capability to predict age and gender in PD multi-lingual corpora.

The extraction of demographic attributes such as gender and age from speech intersects with broader concerns about bias and fairness. A recent study has highlighted that large language models (LLMs) can inadvertently encode and amplify gender biases, particularly when trained on large-scale, uncurated data sources (Xiang, 2026). These concerns extend beyond text-based models and are increasingly relevant for speech and voice technologies, where demographic cues are inherently present in the signal. Another recent study on fairness and privacy in voice biometrics demonstrated that gender has a measurable impact on model behavior when wav2vec 2.0 representations are used for speaker-related tasks (Chouchane et al., 2023). Beyond speech and foundation models, gender differences have been extensively documented in human behavior and health outcomes. For example, behavioral studies conducted after the COVID-19 pandemic revealed systematic gender differences in online purchasing behavior, particularly in the acquisition of protective products, indicating that gender influences decision-making even in large-scale digital environments (Qian et al., 2025). Similarly, epidemiological research has consistently shown sex- and gender-based disparities in COVID-19 severity and mortality, with measurable differences in risk profiles across populations (Díaz-Rodríguez et al., 2023).

In parallel with advances in self-supervised models for general speech and language, the field has seen the emergence of medical domain-adapted foundation models that have come to prominence in late 2025 and early 2026. Notable examples include MedASR, a domain-tuned medical speech-to-text model released by Google Health AI, which achieved substantially lower error rates on clinical dictation and structured medical speech compared with general-purpose automatic speech recognition models. Although most of these initiatives are about to have extensive peer-reviewed evaluations (as of April 2026), they illustrate an important shift toward specialized AI systems in healthcare, with domain-specific speech capabilities increasingly recognized as critical.

From a broader perspective, data-driven phenotyping in neurodegeneration often requires signals that are not easily recoverable from conventional sources such as billing records, medication history, or diagnostic coding alone. Many clinically relevant attributes, disease severity, cognitive status, motor symptoms, comorbid conditions, age-related decline, or progression trajectories, are either under-reported, delayed in documentation, or not represented at all in administrative datasets. This motivates the use of alternative modalities such as speech, which may provide auxiliary structured information and capture subtle but biologically meaningful variation beyond what is typically available in routinely collected clinical data.

Herein, we investigate how accurately a state-of-the-art SFM can estimate gender and age from voice and speech diagnostic-oriented recordings. We evaluate the performance across three corpora that differ in language, task design, and the presence of PD, parkinsonism or related disorders.

## 3. Methods

### 3.1. Datasets

The study employed three independent multilingual datasets comprising a total of 244 unique subjects, including HC, PD patients, and related parkinsonian syndromes.

#### PC-GITA Dataset-1

The PC-GITA dataset (“New Spanish speech corpus database for the analysis of people suffering from Parkinson’s disease”, 2014) consists of speech recordings from 50 HC subjects and 50 individuals with PD, all Spanish speakers from Colombia. The dataset is fully balanced with respect to age and gender. For the PD group, the mean age was 61 ± 9.4 years. For the HC group, the mean age was 61 ± 9.5 years. For our analysis, we used the recordings of read text, the diadochokinetic task “pataka”, and the sustained vowel /a/. Monologue recordings were excluded mostly due to its close similarity with read text.

#### Italian Dataset-2

The Italian dataset (Dimauro et al., 2017) includes recordings from three groups: Young HC (YHC), Elderly HC (EHC), and PD patients. The original dataset contains 15 YHC participants, 22 EHC participants, and 28 PD participants. For the PD group, the mean age was 67.2 ± 8.7 years (9 females, 19 males). For the YHC group, the mean age was 20.8 ± 2.7 years (2 females, 13 males). For the EHC group, the mean age was 67.6 ± 5.3 years (11 females, 7 males). For our analysis, we focused on the read text recordings. For one EHC subject, the read-text sample could not be located (neither the B1-prefixed nor the B2-prefixed files were available); therefore, we used that subject’s word-list recording (prefix PR) instead. For an additional four EHC subjects, we were unable to reliably match their recordings to the corresponding age and gender metadata, and these samples were consequently excluded from the analysis.

#### PD and parkinsonism Dataset-3

To demonstrate possible generalization of the presented solution beyond PD, we employed the dataset described in (Hlavnicka et al., 2019). This dataset contains synthesized replicas of sustained vowels produced by 22 patients with PD, 21 patients with multiple system atrophy (MSA), 18 patients with progressive supranuclear palsy (PSP), and 22 HC. Age and gender distributions are summarized in Section 4.3, Table 5. For consistency, we restricted our analysis to a single copy of the prolonged vowel /A/.

### 3.2. Pre-trained speech models

#### 3.2.1. Preliminaries

In this section, we review some preliminaries about Wav2Vec 2.0 and relevant speech representation learning. Wav2Vec 2.0 is a self-supervised model that learns speech representations directly from raw audio waveforms (Baevski et al., 2020). Let *x* denote a raw speech recording. The model first applies a convolutional encoder that transforms the waveform into a sequence of latent acoustic vectors, *z* = (*z*_1_, …, *z_T_*), where each *z_t_* represents a brief segment of speech. These vectors summarize local acoustic patterns, such as spectral shape and energy, while reducing the length of the signal.

To encourage the model to understand speech in context, some of these acoustic vectors are randomly hidden (masked). The partially masked sequence is then processed by a Transformer network, which produces a new sequence of contextual representations, *c* = (*c*_1_, …, *c_T_*). Each vector *c_t_* combines information from the entire recording, allowing the model to capture longer-term speech structure rather than isolated sounds.

The training objective is based on an idea that the model should be able to infer the missing speech content from its surrounding context. To define what should be predicted, the original acoustic vectors are converted into discrete reference vectors using a quantization step, *q_t_* (discrete target for time step *t*). For each masked position, the model compares its contextual representation *c_t_* with several candidate targets and is rewarded when it assigns the highest similarity to the correct one. In practice, this is implemented using a contrastive loss, which increases the similarity between *c_t_* and the correct target *q_t_*, while decreasing similarity to incorrect alternatives: *correct similarity* ↑, *incorrect similarity* ↓. Similarity between vectors is measured using cosine similarity, 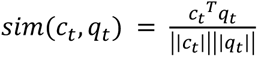. This learning signal forces the model to encode meaningful information about speech content and structure. The contrastive loss at position *t* is typically defined as: 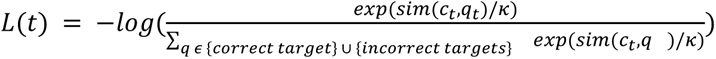, where *κ* is a temperature parameter that controls how strongly the model focuses on the most similar candidates.

As a result of this self-supervised training process, Wav2Vec 2.0 learns contextual speech representations that capture phonetic patterns, speaker traits, and other paralinguistic characteristics, even though no labels are used. These representations form the basis for efficient adaptation to downstream tasks such as age and gender prediction. An illustrative schema of the proposed methodology in this work is presented in Fig. 1.

**Fig. 1.**
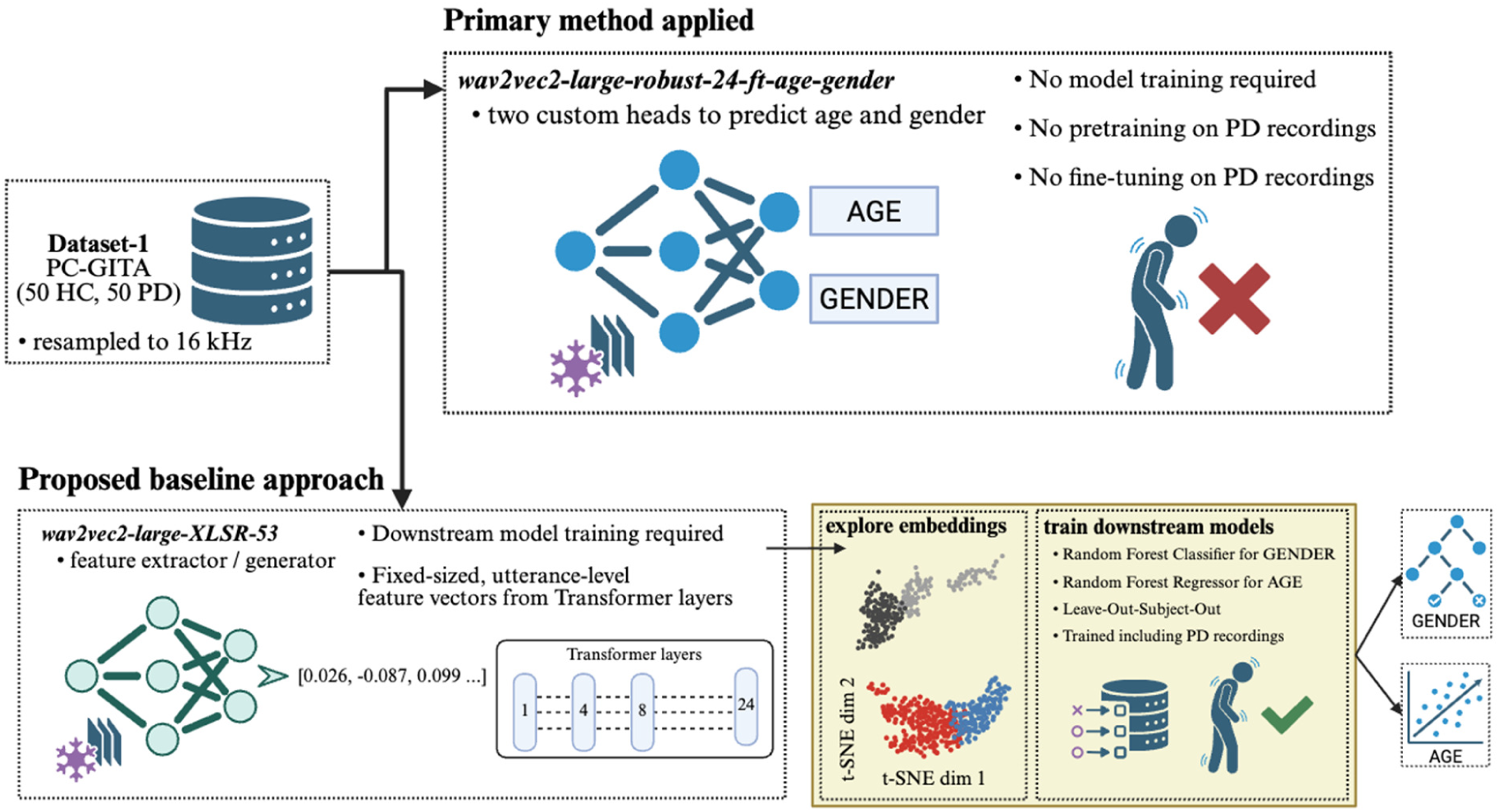
Illustrative schema of the proposed methodology applied to Dataset-1 (Spanish PC-GITA). Wav2Vec 2.0 can be adapted to downstream tasks in different ways. The figure highlights the conceptual difference between (i) fine-tuned model optimized for age and gender prediction, and (ii) the proposed baseline approach, where a pretrained Wav2Vec 2.0 XLSR model was used purely as a feature extractor, followed by training of downstream models. While the fine-tuned model was optimized for age and gender estimation and it is expected that it did not observe PD recordings during training, the downstream classifiers/regressors were trained directly on PD speech data. Overall, the two strategies reflect different trade-offs. Fine-tuning allows the model to adapt fully to the target task, while feature extraction leverages the rich, multilingual representations learned during large-scale self-supervised training.

#### 3.2.2. Model for age and gender recognition based on Wav2vec 2.0

To estimate speaker age and gender, directly without any need to train a downstream model, we employed the 24-layer Wav2Vec 2.0 model for age and gender recognition (Burkhardt et al., 2023). The model takes raw audio waveforms as input and produces continuous age predictions (normalized between 0 and 1, approximately corresponding to 0–100 years) and gender probabilities across three classes: child, female, and male. Because no downstream predictive model was trained (i.e., predictions were obtained directly from the pretrained model without additional optimization step), no train/test data split was required.

This model was obtained by fine-tuning Wav2Vec2-Large-Robust, originally pretrained on 16 kHz speech audio, using several publicly available corpora, including aGender, Mozilla Common Voice, TIMIT, and VoxCeleb2. We accessed the implementation via Hugging Face (Model for Age and Gender Recognition based on Wav2vec 2.0 (24 layers) at Hugging Face, 2023). Given the composition of the pretraining corpora, it is reasonable to assume that the model was not exposed to any neurological-disorder speech datasets during training. For gender inference, we selected the class with the highest predicted probability. All analyses were performed offline with a reasonable CPU runtime, requiring approximately one hour to process a single corpus and no specialized GPU hardware.

#### 3.2.3. Baseline feature-extraction approach using Wav2Vec 2.0 XLSR

To establish a transparent baseline, we employed the Wav2Vec 2.0 XLSR-53 multilingual model (Xu et al., 2021; Wav2Vec2-XLSR-53, 2020) as a fixed feature extractor. This choice reflects a commonly adopted strategy in biomedical speech analysis, where pretrained speech models are used to generate representations without task-specific fine-tuning. Hidden representations were extracted from multiple Transformer layers (layers 1, 4, 8, and the final layer 24) to capture information at different levels of abstraction. For each selected layer, frame-level features were temporally aggregated using mean pooling, resulting in layer-specific utterance-level embeddings with a fixed dimensionality of 1024.

These embeddings served as input to lightweight downstream models for demographic prediction. Gender classification was performed using a Random Forest classifier with n = 80 estimators, while age estimation was addressed using a Random Forest regressor with the same number of estimators. No hyperparameter optimization was conducted. To obtain subject-level predictions and ensure a fair comparison with the proposed approach, we adopted a leave-one-subject-out cross-validation (LOSO-CV) strategy. This evaluation protocol enabled unbiased assessment at the participant level and allowed direct comparison with the main model’s subject-level performance.

Finally, to qualitatively assess the extent to which demographic information is inherently encoded in the extracted representations, we visualized the utterance embeddings using t-distributed stochastic neighbor embedding (t-SNE) with a perplexity of 30. These visualizations provide additional insight into the separability of demographic attributes within the pretrained feature space.

### 3.3. Age and gender prediction

Each speech recording was paired with the participant’s chronological age “true age” and the model-estimated age “predicted age”, with corresponding gender attributes following the same principle. To facilitate comparison of the overall age distributions, both true and predicted ages were discretized into 10-year bins: <30, 30–39, 40–49, 50–59, 60–69, 70–79, and ≥80 years. The prediction performance was evaluated using label agreement between the participant’s true value and the model-predicted value.

### 3.4. Statistical methods

Beyond reporting simple accuracy metrics, the primary objective of the statistical analysis was to determine whether the age-prediction performance in individuals with PD was comparable to that observed in HC. Demonstrating equivalence or non-inferiority of predictive performance in PD would support the applicability of the model in clinical or research settings involving this population. For all statistical tests, a two-sided p-value < 0.05 was considered indicative of statistical significance.

For each defined group, we compared the predicted and true age distributions across the predefined 10-year bins. The Chi-square statistic quantified the deviation between observed (true) and model-estimated bin frequencies, treating the true age distribution as the reference. This test evaluated whether the model’s predictions preserved the overall age structure within each group. In addition, to assess rank-order agreement and overall predictive validity at the individual level, we computed Spearman’s rank correlation coefficient with its corresponding p-value between true and predicted continuous ages.

Agreement between true and predicted age was further evaluated using Bland–Altman plots, reporting mean bias and 95% limits of agreement, more details can be found in Supplementary Methods.

## 4. Results

Unless stated otherwise, the results presented in this section correspond to the model described in Section 3.2.2., which constitutes the primary model of interest in this study. To enable a direct and fair comparison with the proposed baseline approach, comparative evaluations are reported exclusively on Dataset-1 (Section 4.1.1). Results obtained on the remaining datasets are reported separately and are used primarily to assess generalization and robustness, rather than baseline comparability.

### 4.1. Dataset-1

The achieved results of gender and age estimation for the Spanish PC-GITA dataset are summarized in Table 1. Gender classification accuracy reached consistently high values (min. 94%) across all speech tasks (read text, pataka, sustained vowel /a/) and for both studied groups, i.e., HC and PD. Specifically, for the read text task, perfect classification accuracy was achieved for the HC group (100%), while accuracy reached 98% for the PD group.

**Table 1.**
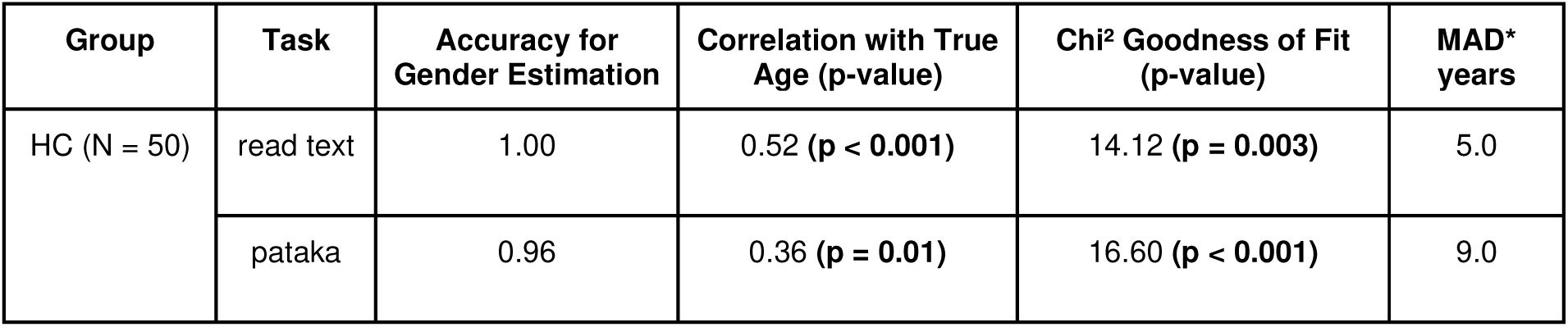
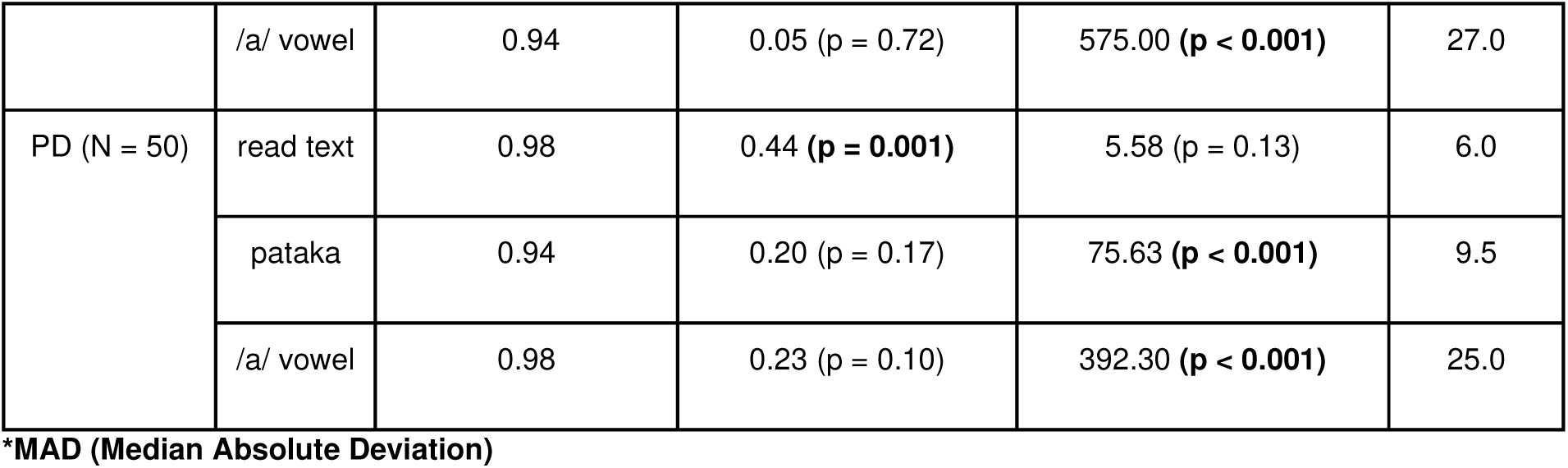
Gender classification accuracy and age estimation results for the Spanish PC-GITA dataset across speech tasks and participant groups. Age was discretized into categories {<50, 50–59, 60–69, 70+}. Since the <30, 80–89, and 90+ age bins contained no observed samples, they were merged with adjacent categories to avoid zero expected cell counts in the chi-square tests.

| Group | Task | Accuracy for Gender Estimation | Correlation with True Age (p-value) | Chi<sup>2</sup> Goodness of Fit (p-value) | MAD* years |
| --- | --- | --- | --- | --- | --- |
| HC (N = 50) | read text | 1.00 | 0.52 ($p < 0.001$) | 14.12 ($p = 0.003$) | 5.0 |
| | pataka | 0.96 | 0.36 ($p = 0.01$) | 16.60 ($p < 0.001$) | 9.0 |
|  | /a/ vowel | 0.94 | 0.05 (p = 0.72) | 575.00 (p < 0.001) | 27.0 |
| PD (N = 50) | read text | 0.98 | 0.44 (p = 0.001) | 5.58 (p = 0.13) | 6.0 |
|  | pataka | 0.94 | 0.20 (p = 0.17) | 75.63 (p < 0.001) | 9.5 |
|  | /a/ vowel | 0.98 | 0.23 (p = 0.10) | 392.30 (p < 0.001) | 25.0 |

\*MAD (Median Absolute Deviation)

Regarding age estimation, the principal findings indicate statistically significant correlations between the predicted and true age for the read text task in both groups. For HC, a moderate positive correlation was observed (Spearman’s ρ = 0.52, p < 0.001), while for PD a slightly weaker but still significant correlation was obtained (ρ = 0.44, p = 0.001). These results demonstrate a statistically significant ability of the foundation model to estimate age, even in speakers affected by PD. In addition, chi-square goodness-of-fit tests were applied to compare predicted and true age distributions. In the majority of cases, the tests indicated statistically significant differences between the distributions. Only one non-significant result was observed, namely for the read text task in the PD group (χ² = 5.58, p = 0.13), suggesting no substantial violation of the estimated age distribution in this group.

To further assess the agreement between predicted and true ages at both the individual and category levels, scatter plots (Fig. 2) and boxplots (Fig. 3) were generated for the HC group. Corresponding visualizations for the PD group are provided in Fig. 4 and Fig. 5. These figures reveal that while overall age-related trends were reasonably well captured for the read text and pataka tasks, the sustained vowel task failed to reflect the true age distribution. In this task, a pronounced underestimation of age was observed for both HC and PD groups, indicating a clear failure of the model in capturing age-related information from vowel phonation alone. Age-prediction agreement in the HC and PD cohorts was further examined using Bland–Altman analysis (Supplementary Figs. S1 and S2).

**Fig. 2.**
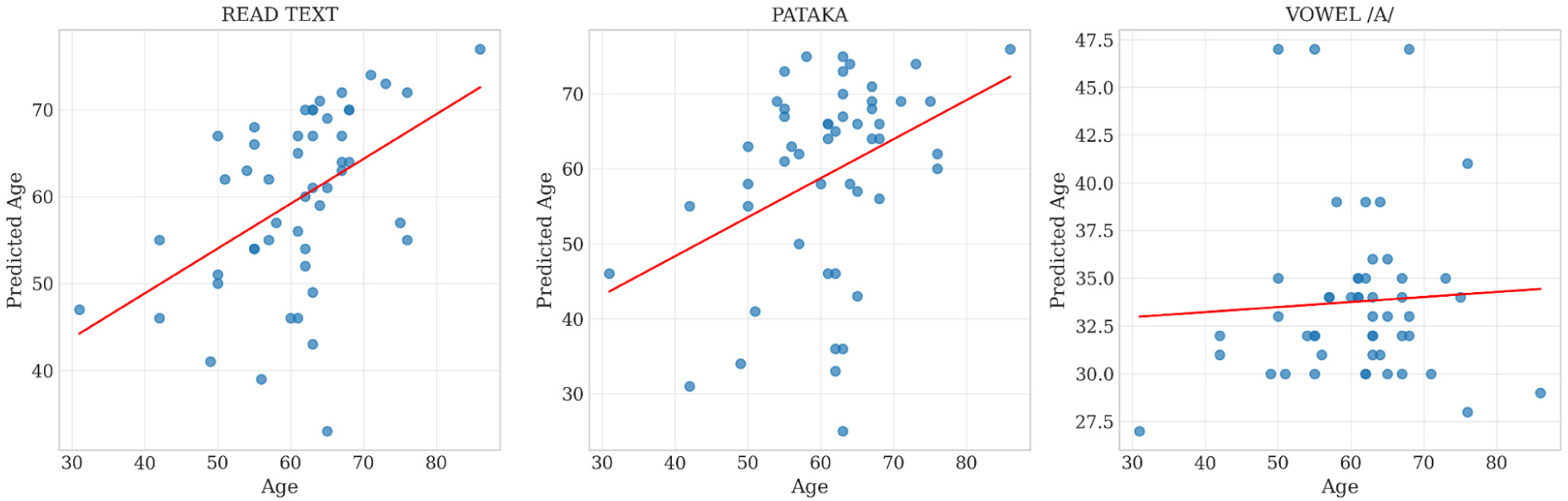
Scatter plots showing the relationship between true age and predicted age for the HC group across the three speech tasks: read text, pataka, and sustained vowel (/a/).

**Fig. 3.**
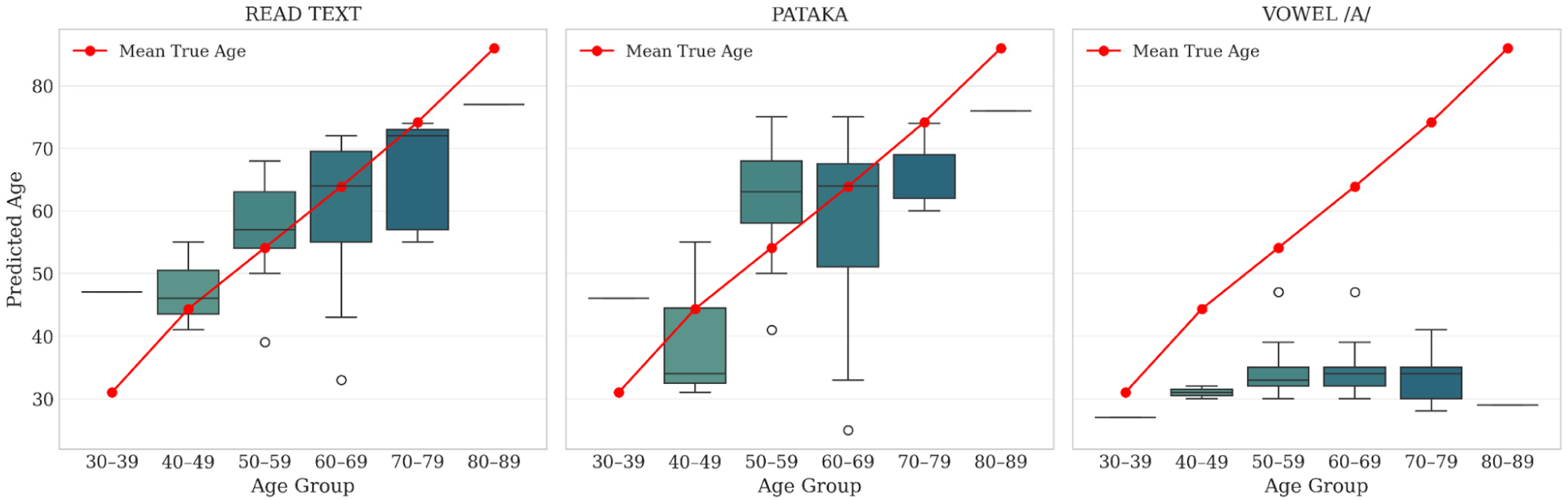
Boxplots of predicted age across true age groups for the HC group. The mean true age for each group is overlaid as a reference marker.

**Fig. 4.**
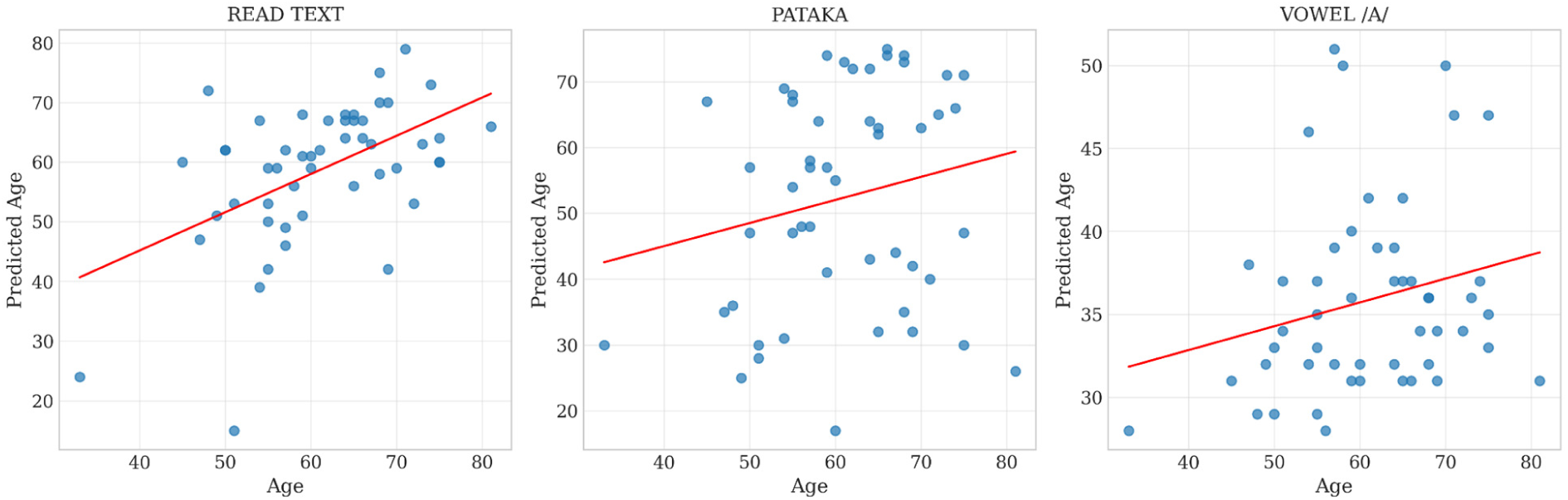
Scatter plots showing the relationship between true age and predicted age for the PD group across the three speech tasks: read text, pataka, and sustained vowel (/a/).

**Fig. 5.**
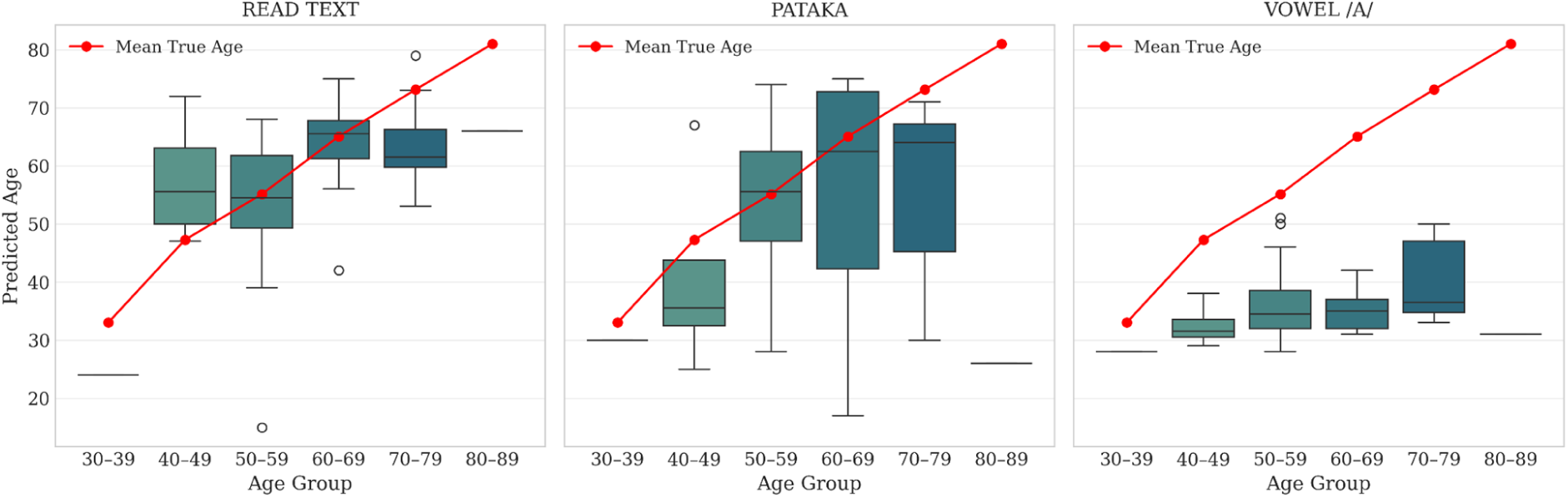
Boxplots of predicted age across true age groups for the PD group. The mean true age for each group is overlaid as a reference marker.

#### 4.1.1. Comparisons with the proposed Wav2Vec 2.0 baseline

Dataset-1 was selected for this purpose because it provided the largest sample size for both HC and PD subjects among the available datasets. In addition, Dataset-1 included three distinct diagnostic-oriented speech recording types, making it particularly well suited for evaluating task-dependent model behavior and demographic metadata estimation. The baseline comparison focused on two speech tasks, read text and sustained vowel phonation.

Across both tasks and subject groups, deeper layers generally yielded better performance (Table 2). This trend was consistent for both HC and PD groups. In the case of gender classification, performance degradation of the proposed baseline was observed across both speech tasks. For the read-text task, accuracy dropped by 8% for HC and 12% for PD, even when using the best-performing layer (first Transformer layer). Similarly, for the vowel phonation task, gender classification accuracy decreased by approximately 8% for both HC and PD in the best-performing layer. For age estimation, the baseline approach exhibited clear limitations. Performance deteriorated not only for the vowel phonation task, but also for the read-text task, and this degradation was observed consistently across both HC and PD groups.

**Table 2.** Baseline performance across Transformer layers. Principal Component Analysis (95% variance retained) was applied for age estimation.

| Group | Task | Layer | Accuracy for Gender Estimation | Correlation with True Age |
| --- | --- | --- | --- | --- |
| HC (N = 50) | read text | 1T | 0.92 | -0.32 |
|  |  | 4T | 0.82 | 0.10 |
|  |  | 8T | 0.76 | 0.06 |
|  |  | LH | 0.70 | 0.12 |
|  | /a/ vowel | 1T | 0.86 | 0.08 |
|  |  | 4T | 0.86 | 0.02 |
|  |  | 8T | 0.80 | -0.06 |
|  |  | LH | 0.70 | -0.08 |
| PD (N = 50) | read text | 1T | 0.86 | 0.17 |
|  |  | 4T | 0.84 | -0.12 |
|  |  | 8T | 0.78 | -0.22 |
|  |  | LH | 0.71 | 0.03 |
|  | /a/ vowel | 1T | 0.90 | -0.35 |
|  |  | 4T | 0.78 | 0.10 |
|  |  | 8T | 0.78 | 0.20 |
|  |  | LH | 0.72 | 0.06 |

To further investigate whether demographic information is inherently encoded in the pretrained embeddings, we visualized the utterance-level representations using t-SNE. For gender, a promising degree of separability was observed across multiple layers (Fig. 6 and Fig. 7), indicating that gender-related information is consistently embedded in the representation space. In contrast, for age, no clear visual clustering or separability was evident in the t-SNE projections (Fig. 8 and Fig. 9), supporting the quantitative findings and suggesting that age-related information is weakly or inconsistently encoded.

**Fig. 6.**
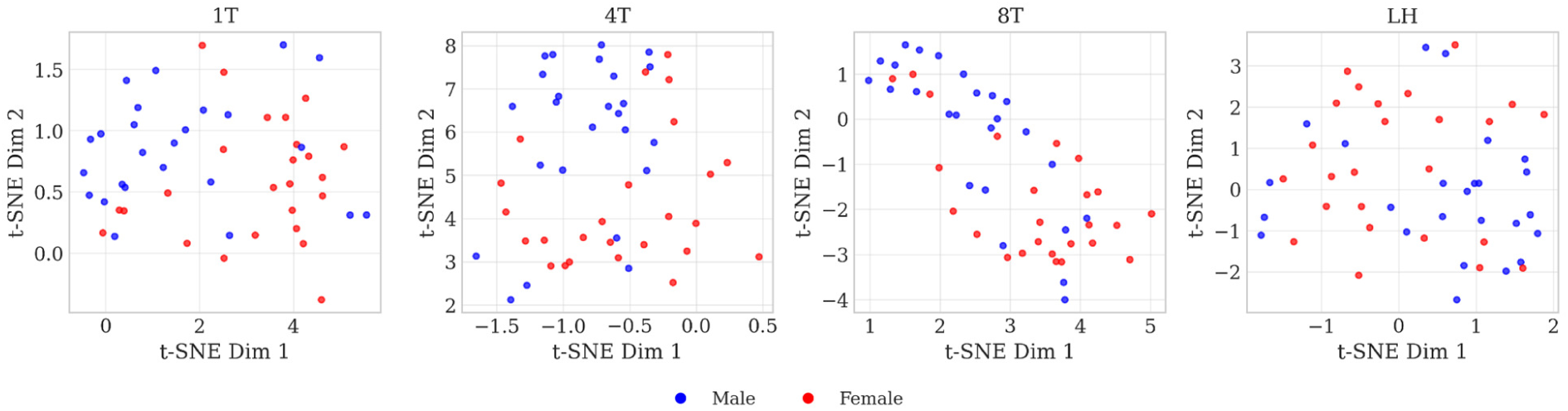
t-SNE visualization of Wav2Vec 2.0 embeddings extracted from read-text speech of HC, illustrating separability with respect to gender.

**Fig. 7.**
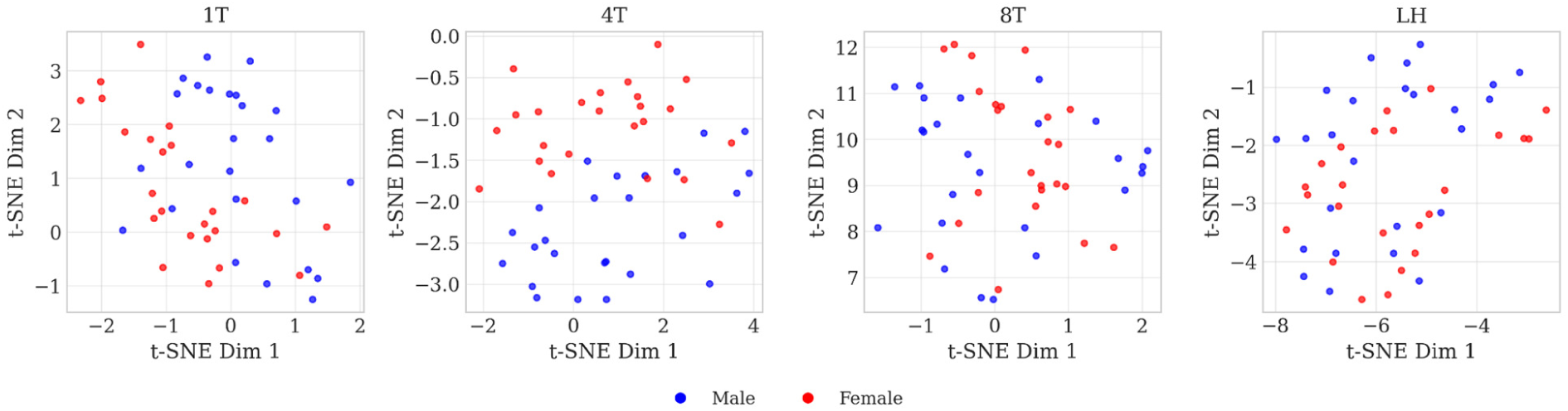
t-SNE visualization of Wav2Vec 2.0 embeddings extracted from read-text speech of PD, illustrating separability with respect to gender.

**Fig. 8.**
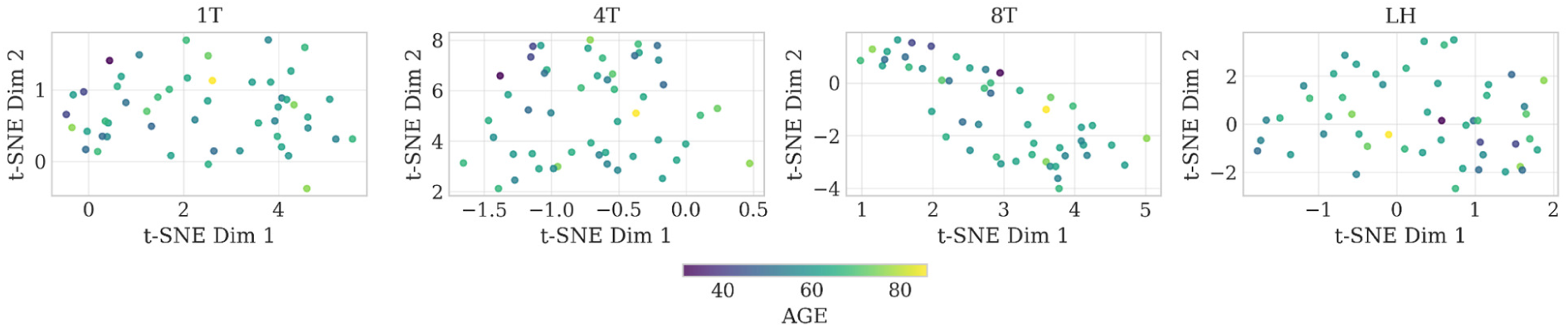
t-SNE visualization of Wav2Vec 2.0 embeddings extracted from read-text speech of HC, colored by age, showing no clear age-related clustering.

**Fig. 9.**
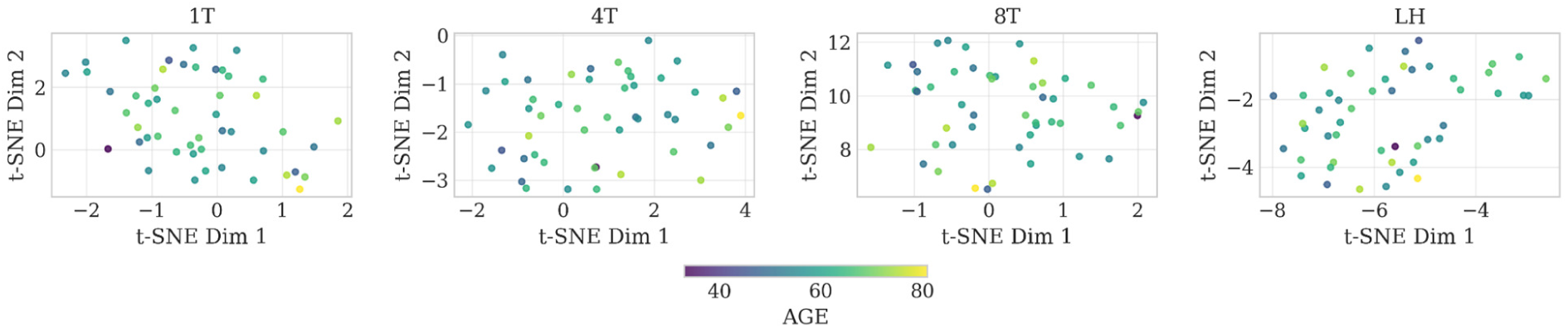
t-SNE visualization of Wav2Vec 2.0 embeddings extracted from read-text speech of PD, colored by age, showing no clear age-related clustering.

#### 4.1.2. Exploratory LLM-based validation

To contextualize errors observed in automatic age prediction from sustained vowels, we conducted a small LLM-based exploratory analysis. As shown in Fig. 5, we observed substantial prediction errors in age estimation from sustained vowel phonations, suggesting that age-related information may be weakly encoded or confounded in this speech mode. Given recent advances in multimodal LLMs, we explored whether an LLM-based listening paradigm might offer complementary cues that are not captured by purely acoustic self-supervised representations.

A small subset of three recordings was selected to showcase this idea. Each recording consisted of short clinical speech material (including two recordings of the sustained vowel /a/ and one recording of pataka). The recordings were presented individually to ChatGPT (GPT-5.2; December 2025), which was instructed to provide a single numeric age estimate per recording, treat the estimate as approximate and heuristic, and avoid claims of precision or clinical validity. No speaker metadata (including true age) was provided to the model. The resulting LLM-based estimates were then compared post hoc (Table 3) with both the true chronological age and the age predicted by the Wav2Vec 2.0 model (Section 3.2.2). The LLM-based estimates were observed to be closer to the true age range in two examples of the sustained vowel /a/, but did not improve the estimation in the case of pataka. This observation highlights a potentially promising direction for future research aimed at mitigating or addressing the current limitations. Despite the promise of multimodal LLMs, SFMs currently remain the preferred choice, as they provide fixed, reproducible embeddings that enable controlled experiments with audio recordings.

**Table 3.**
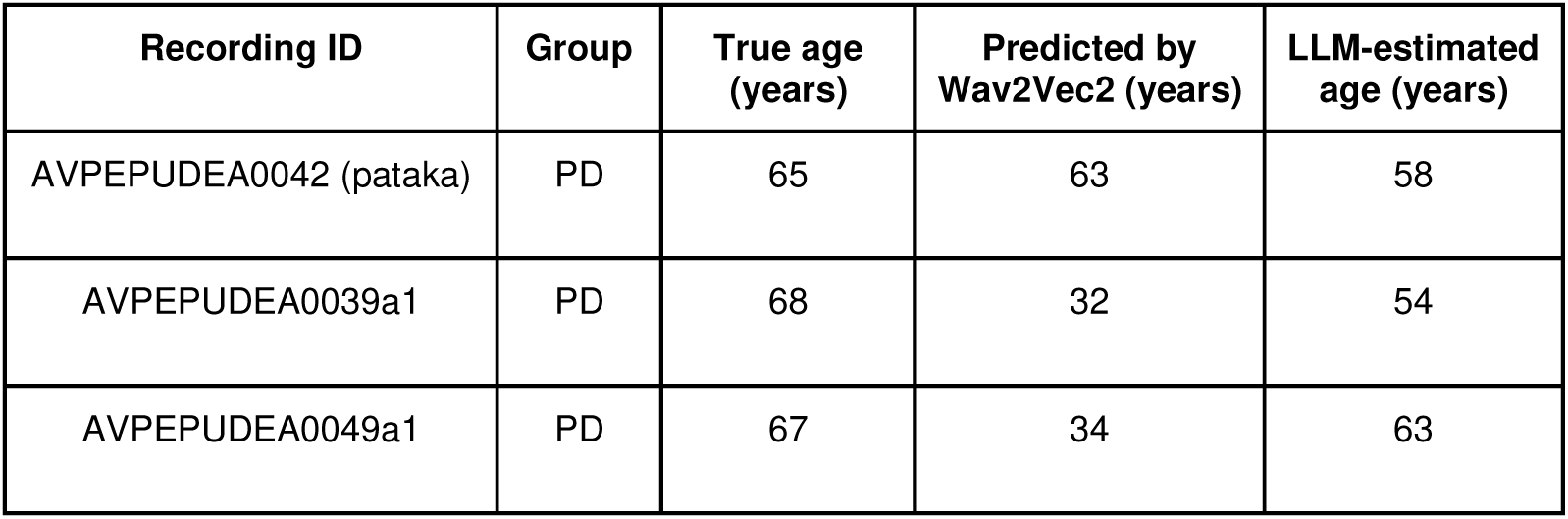
Summary of the LLM-based showcase. GPT-5.2 showed promise to improve performance of estimating age from sustained vowels /a/ recordings.

| Recording ID | Group | True age (years) | Predicted by Wav2Vec2 (years) | LLM-estimated age (years) |
| --- | --- | --- | --- | --- |
| AVPEPUDEA0042 (pataka) | PD | 65 | 63 | 58 |
| AVPEPUDEA0039a1 | PD | 68 | 32 | 54 |
| AVPEPUDEA0049a1 | PD | 67 | 34 | 63 |

### 4.2. Dataset-2

The results for the Italian dataset are summarized in Table 4. Compared to the Spanish PC-GITA dataset, this dataset is characterized by a smaller sample size and a different group structure, comprising YHC, EHC, and PD. The analysis was limited to the read text task. Gender estimation achieved perfect classification accuracy (100%) across all three groups.

**Table 4.**
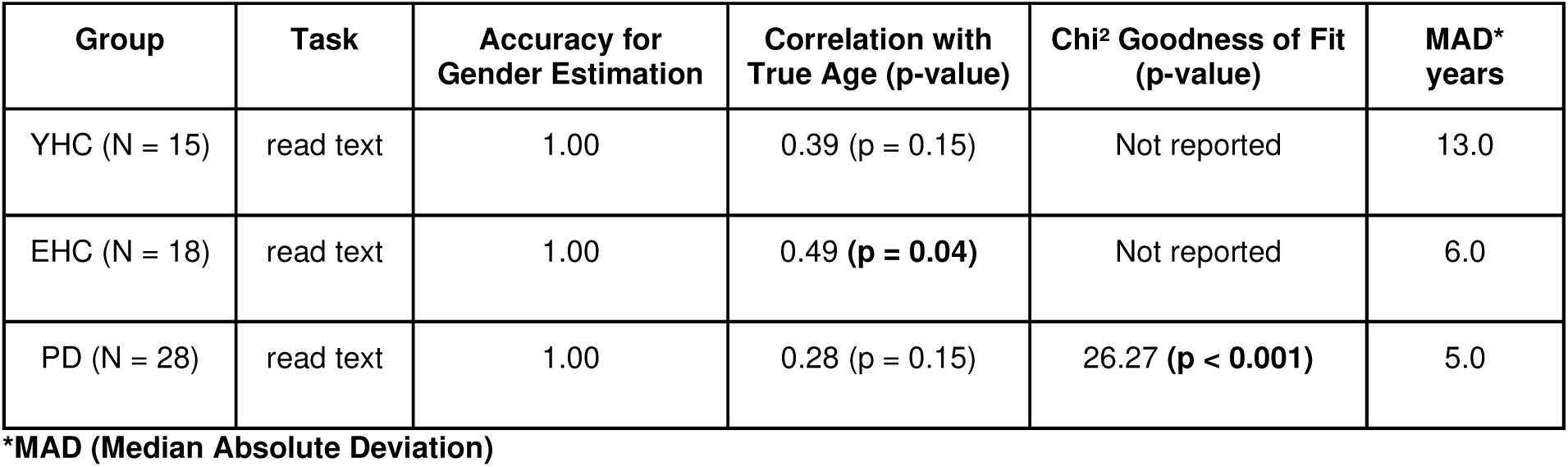
Gender classification accuracy and age estimation results for the Italian dataset (read text task only) across YHC, EHC, and patients with PD.

| Group | Task | Accuracy for Gender Estimation | Correlation with True Age (p-value) | Chi<sup>2</sup> Goodness of Fit (p-value) | MAD* years |
| --- | --- | --- | --- | --- | --- |
| YHC (N = 15) | read text | 1.00 | 0.39 ($p = 0.15$) | Not reported | 13.0 |
| EHC (N = 18) | read text | 1.00 | 0.49 (**$p = 0.04$**) | Not reported | 6.0 |
| PD (N = 28) | read text | 1.00 | 0.28 ($p = 0.15$) | 26.27 (**$p < 0.001$**) | 5.0 |

\*MAD (Median Absolute Deviation)

With respect to age estimation, moderate positive correlations between predicted and true age were observed, although statistical significance varied across groups. For the EHC group, the correlation reached statistical significance (Spearman’s ρ = 0.49, p = 0.04), whereas for YHC (ρ = 0.39, p = 0.15) and PD (ρ = 0.28, p = 0.15) the correlations did not reach significance. Chi-square goodness-of-fit testing was performed only for the PD group, where a statistically significant difference between predicted and true age distributions was observed (χ² = 26.27, p < 0.001). For the YHC and EHC groups, chi-square tests were not reported due to insufficient cell counts required to satisfy test assumptions. To visually assess model performance, scatter plots illustrating the relationship between true and predicted age for the read text task are presented in Fig. 10 for YHC, EHC, and PD groups. Complementary boxplots are shown in Fig. 11. Age-prediction bias across YHC, EHC, and PD groups was assessed using Bland–Altman analysis (Supplementary Fig. S3).

**Fig. 10.**
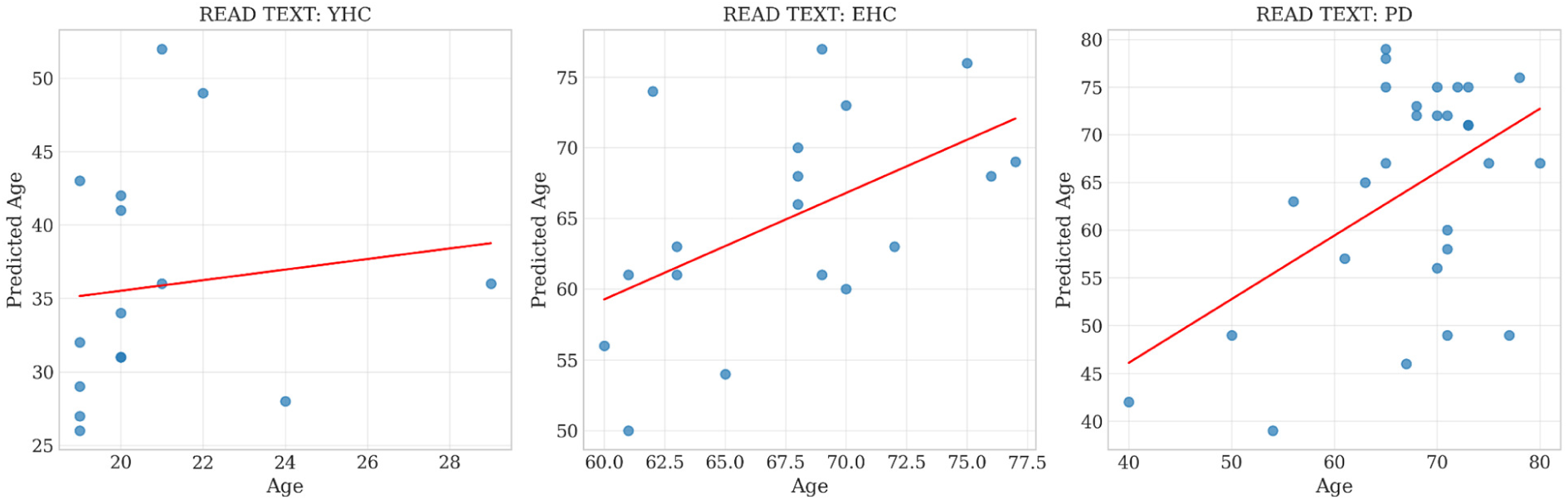
Scatter plots illustrating the relationship between true age and predicted age for the read text task in the Italian dataset.

**Fig. 11.**
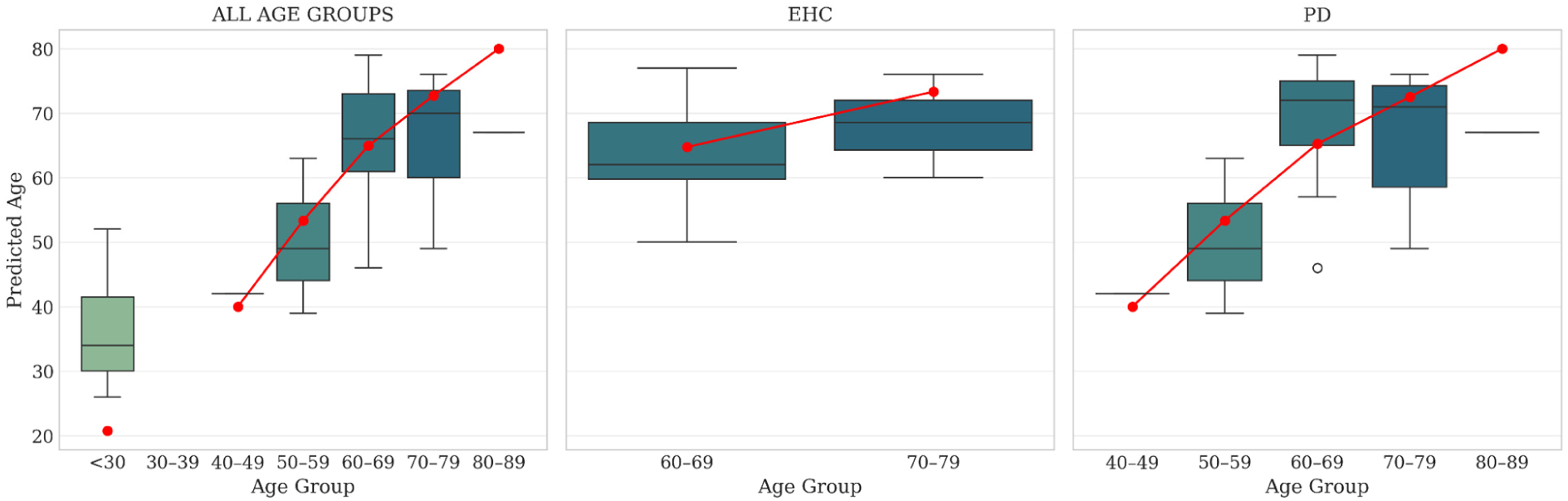
Boxplots of predicted age for the Italian dataset based on the read text task. The leftmost plot shows results across all age groups, followed by separate visualizations for the EHC and PD. The YHC group is not shown due to its narrow age range. Notably, the 50–79 age interval was well captured by the model (typical for PD onset).

### 4.3. Dataset-3

To assess the potential generalization of the proposed approach beyond PD, we evaluated its performance on Dataset-3, which included synthesized sustained vowel productions from patients with PD, MSA, PSP and HC. Unlike Datasets 1 and 2, individual-level ground-truth annotations were not available for this dataset. Therefore, the evaluation was performed at the group level, and the reported metrics followed a different format.

Gender and age estimation performances across diagnostic groups are summarized in Table 5. A chi-square goodness-of-fit test comparing true and predicted gender distributions across all groups revealed no statistically significant differences (χ² = 2.2, df = 7, p = 0.95), indicating that the predicted gender proportions matched the true distributions at the group level. In contrast, age estimation showed a pronounced systematic underestimation across all diagnostic categories. While the true average ages ranged between approximately 61 and 67 years, the predicted average ages were substantially lower, spanning from 29.8 to 36.1 years. This underestimation was consistent across HC, PD, MSA, and PSP groups.

**Table 5.** Group-level comparison of true and predicted gender counts and average age estimates, including HC, PD, MSA, and PSP. Age values are reported as group means. Due to the absence of individual-level ground-truth annotations, evaluation was performed at the group level only.

| Estimated | F (true/predicted) | M (true/predicted) | avg. age (true/predicted) |
| --- | --- | --- | --- |
| HC (N = 22) | 11 / 11 | 11 / 11 | 63.6 / 29.8 |
| MSA (N = 21) | 12 / 16 | 9 / 5 | 61.0 / 36.1 |
| PD (N = 22) | 12 / 14 | 10 / 8 | 64.4 / 34.2 |
| PSP (N = 18) | 6 / 5 | 12 / 13 | 66.7 / 32.2 |

## 5. Discussion

True value of the foundational models lies in understanding how existing pretrained models behave when applied to clinically relevant problems where prior evidence is limited. In this context, the present study should be viewed primarily as a validation study in a domain with possibly missing or limited knowledge, rather than as the development of a novel algorithm.

In this study, we systematically evaluated the pretrained speech foundation model, Wav2Vec 2.0, across multiple datasets, speech tasks, and neurological conditions, including PD and related parkinsonian syndromes. This design enabled a realistic assessment of what can (and cannot) be expected from such a model when deployed without task-specific training. Our findings demonstrated near-perfect performance in gender estimation across all datasets and speech tasks, confirming that gender-related information is robustly encoded in pretrained speech representations. While age estimation proved more challenging, the model consistently captured meaningful age-related structure, allowing reliable characterization of datasets and disease-specific age distributions in both HC and patients.

Importantly, unlike the previous work on age prediction using Wav2Vec-based models (Klempíř & Krupička, 2024), with employing the wav2vec2-large-robust-24-ft-age-gender (3.2.2), no training or fine-tuning was required in this study. Our objective was not to maximize predictive performance but to assess the intrinsic capability of a general pretrained model. This constraint can be viewed as a methodological strength, i.e. by preventing any model from “seeing” the evaluated data during training, we reduce the risk of overfitting and provide a clearer picture of true model generalization. Moreover, using the model documented in (Wav2Vec2-XLSR-53, 2020), both age and gender can be estimated in a single execution, eliminating the need to deploy separate task-specific models. According to the model card documentation, it is unlikely that the evaluated foundation model was exposed to dysarthric speech during pretraining, yet it still achieved reasonable accuracy within the PD subgroup. This observation suggests that task-specific fine-tuning may not be strictly required for reliable gender estimation in pathological speech. Based on the results observed for Datasets 1 and 2, a broader representation of older speakers (typically 80+ years; undetected) in pretraining data may further contribute to improved age prediction performance.

Quantitatively, analyses on Datasets 1 and 2 did not reveal a systematically higher age prediction error in the patient subgroups. In PD speakers from the PC-GITA dataset, the MAD was 6.0 years, compared to 5.0 years in HC. Similarly, in the Italian dataset, the MAD was 6.0 years for EHC and 5.0 years for PD subjects. MAD is a commonly used metric for evaluating age prediction from voice acoustics, e.g. (Novotny et al., 2023). In (Novotny et al., 2023), focused on predicting children’s age and achieved precise estimates even using sustained vowels, albeit with specialized algorithms, not foundation models.

For Dataset 1, comparison with the proposed baseline (Section 3.2.3) showed that a model trained on task-specific embeddings could not achieve performance comparable to that reported in Section 4.1, even when trained explicitly on PD subjects. In more detail, for age estimation, the baseline failed not only on sustained vowel phonation but also on read-text tasks, and this trend was consistent across both HC and PD participants. To place these findings in context, (Klempíř & Krupička, 2024) reported that wav2vec embeddings could capture age-related structure with a Spearman correlation of ρ = 0.59. However, their dataset included only two clearly separable age groups, which simplified the prediction task. By contrast, the PC-GITA dataset covers a broader age range (approximately 40+ years).

As observed in Datasets 1 and 3, sustained vowel phonation (/a/) consistently led to substantial underestimation of age. Importantly, this bias was not only specific to HC and PD (Table 1; HC MAD = 27.0; PD MAD = 25.0; Bland–Altman Supplementary Figs. S1 and S2) but was also present in atypical parkinsonian syndromes (Table 5), indicating a generally limited age-discriminative capacity of the model when restricted to isolated vowel production. Nevertheless, the accurate preservation of gender distributions across all diagnostic categories suggests that certain speaker characteristics remain robustly encoded even under these constrained conditions. The systematic failure of age estimation from sustained vowels may have implications for downstream representation learning. Specifically, the weak age sensitivity observed for vowel-only recordings may be advantageous when using Wav2Vec-based embeddings for classification tasks, as it suggests that age-related variation plays a limited role and is less likely to dominate the learned representations. In contrast, given the consistent and robust performance observed for gender estimation across all tasks and datasets, the use of gender-balanced datasets appears advisable to prevent unintended bias propagation in speech-based modeling pipelines.

We acknowledge that gender recognition from speech might raise important ethical considerations. In this study, our objective is not to infer or enforce personal identity attributes, but rather to distinguish subject subgroups in neurodisordered datasets where systematic differences in speech production are known to exist. Prior research has consistently demonstrated gender-related differences in speech characteristics, including in neurological populations (Cao et al., 2025; Skodda et al., 2011; Tykalova et al., 2025). Capturing such variability is essential for correctly characterizing datasets and avoiding confounding effects in downstream analyses. At the same time, we recognize that some individuals may not wish to disclose or be categorized by gender, which further motivates the need for transparent and responsible use of such annotations. In this context, age and gender stratification strategies, such as training on one subgroup and testing on another, are particularly relevant for studies employing speech machine learning models for condition detection. Such stratified analyses can help disentangle disease-related speech markers from demographic effects, improve interpretability, and provide a more rigorous assessment of model generalization across populations.

Beyond controlled diagnostic-oriented datasets, automated estimation of age and gender may also be valuable in scenarios where demographic information is completely missing or poorly documented, such as speech samples collected from publicly available sources (e.g., online videos or interviews). Recent studies, including ParkCeleb (Favaro et al., 2024) and ADCeleb (Gao et al., 2025), illustrated a promising trend toward leveraging publicly available recordings of individuals who have disclosed neurological conditions, enabling large-scale and, in some cases, longitudinal analyses of speech. As pretraining datasets continue to grow in size and diversity, the use of such data is expected to increase, and existing foundation models may play an important role in the automated annotation and initial characterization of these samples. An additional practical benefit arises in studies where fully balanced datasets are not available. In such cases, reporting other “baseline” performance metrics, beyond straightforward sample-size baselines, can provide valuable context.

A pretrained model can be useful not only in cases where age and gender information is missing. Having access to a general-purpose, large pretrained model for age and gender estimation that performs reasonably well in PD and other voice or speech disorders offers advantages even when demographic metadata are available. In such settings, the model can serve as an independent reference for identifying potential data quality issues. For example, during the development of speech data pipelines, recordings may be incorrectly paired, or metadata may be inconsistently assigned to individual samples. Discrepancies between predicted and recorded demographic attributes can thus function as a form of automated self-quality check, highlighting samples that warrant closer inspection.

A known limitation of the current approach is that, when the goal is to achieve the highest possible predictive accuracy, particularly for age estimation, performance is likely to be substantially lower than what could be achieved with task-specific fine-tuning. In such cases, fine-tuning the pretrained model on disorder-specific data, for example PD or other dysarthric speech, would be necessary to improve age prediction accuracy. While our study demonstrated the intrinsic capability of a dataset-agnostic foundation model, we believe that fine-tuning remains the most effective strategy for applications that demand precise demographic predictions.

Future work should investigate multimodal phenotyping strategies combining speech-derived features with records of clinical care or administrative health data. For conditions such as PD or AD, reliably distinguishing demographic structure is only a first step; more complex voice-based signatures of disease severity, motor impairment, or cognitive decline could help address known limitations of structured data sources, where staging and functional trajectories are difficult to recover at scale. Furthermore, future work should investigate whether multimodal LLMs, such as GPT-5.2 that can directly process audio, can provide meaningful clinical information beyond what is captured by self-supervised representations. Direct audio LLMs are evolving quickly and could serve as a future tool for inferring speaker characteristics directly from speech without relying solely on SFMs embeddings, especially for pathological recordings.

## 6. Implications

### 6.1. Theoretical implications

The present study provides empirical evidence from biomedical speech experiments highlighting the importance of task-specific fine-tuning of SFMs. In particular, our results demonstrated that the model specifically designed for age and gender prediction benefited from task-oriented adaptation, rather than relying solely on general-purpose pretraining. Importantly, the findings support the notion that pathological speech does not need to be present in the pretraining corpus for SFMs to successfully capture demographic information. That is, gender can be reliably classified, and age category can be estimated, even when the model has not previously encountered pathological examples. Regarding the limitations in age estimation for certain diagnostic-oriented recordings, particularly sustained vowel phonation, there remains a theoretical opportunity for further task-specific fine-tuning. However, to our knowledge, this potential is constrained by the limited availability of publicly accessible datasets for such specialized speech tasks.

### 6.2. Practical implications

First, beyond the primary focus on pathological recordings, our study demonstrates the utility of SFMs across different diagnostic-oriented speech tasks in HC, indicating their potential applicability in biometric systems. Second, these models can support the identification of demographic information in datasets that are demographically imbalanced or require automated annotation of large-scale speech corpora. Third, independently pretrained SFMs can help identify inconsistencies in speech processing pipelines, for example by verifying whether assigned dataset labels are internally consistent with the underlying speech representations. This, in turn, provides the opportunity to develop additional specialized SFMs focused on predicting clinically and acoustically meaningful parameters, including demographic and other relevant audio-based attributes. SFMs should be understood not only as predictive tools, but as hybrid software–data assets, since the representations they generate can be reused across multiple downstream applications.

## 7. Conclusion

SFMs, such as Wav2Vec 2.0, have already demonstrated strong utility in representation learning, classification, and automatic speech recognition for PD. Here, we extend their application to the annotation of PD speech recordings for demographic estimation, specifically gender and age. Our results showed that gender can be predicted with excellent accuracy across healthy and pathological speakers. Age estimation was less precise, particularly for older individuals, but the model still captures meaningful structure, allowing characterization of datasets in both HC and, importantly, PD patients.

These findings suggest broader potential applications of general pretrained speech models. They can support quality control of recordings in data pipelines for voice disorder research, detect mislabeled samples, and highlight inconsistencies within datasets, serving as exploratory tools without requiring task-specific training. While fine-tuning the model on the target data could improve age estimation, the current approach demonstrates that large pretrained models already provide valuable insights and can characterize datasets reliably with zero/minimal exposure to training data.

## Data Availability

All data produced in the present study are available upon reasonable request to the authors.

## CRediT authorship contribution statement

**Ondrej Klempir**: Writing – original draft, Visualization, Validation, Methodology, Investigation, Formal analysis, Data curation, Conceptualization. **Ales Tichopad**: Writing – review & editing, Formal analysis, Methodology, Funding acquisition. **Radim Krupicka**: Writing – review & editing, Formal analysis, Supervision, Project administration, Funding acquisition. All authors reviewed the results and approved the final version of the manuscript.

## Ethics statements and data availability

The datasets used in this study are publicly available, with one dataset - PC-GITA - being available for download upon request from their authors. The PC-GITA dataset is available upon request from Juan Rafael Orozco-Arroyave affiliated with Universidad de Antioquia UdeA. The study complied with the Helsinki Declaration and was approved by the Ethics Committee of Clinica Noel in Medellín, Colombia. A written informed consent was signed by each participant.

## Role of the funding source

The authors declare that the study sponsors had no role in the design of the study, the collection, analysis, and interpretation of data, the writing of the manuscript, or the decision to submit the manuscript for publication. All sources of funding are duly acknowledged, and no external influence has affected the integrity or independence of this research.

## Declaration of Generative AI and AI-assisted Technologies in Scientific Writing

During the preparation of this work the authors used ChatGPT to improve the readability and language of the manuscript. After using this tool, the authors reviewed and edited the content as needed and take full responsibility for the content of the published article.

## Acknowledgement

This research was supported by the Czech Health Research Council grant no. NW24-04-00259. This research was partially supported by the project CZ.02.01.01/00/23_025/0008743, funded by the European Union under the Operational Programme Johannes Amos Comenius (OP JAK).

## Competing interests

The authors declare no competing interests.

## Notes

### Competing Interest Statement

The authors have declared no competing interest.

### Author Declarations

The datasets used in this study are publicly available, with one dataset being available for download upon request from their authors. 1. (Italian datataset) Available at: https://ieee-dataport.org/open-access/italian-parkinsons-voice-and-speech 2. (PC-GITA) The PC-GITA dataset is available upon request from Juan Rafael Orozco-Arroyave affiliated with Universidad de Antioquia UdeA. The study complied with the Helsinki Declaration and was approved by the Ethics Committee of Clinica Noel in Medellin, Colombia. A written informed consent was signed by each participant. 3. Available at: https://figshare.com/articles/dataset/Synthetic_vowels_of_speakers_with_Parkinson_s_disease_and_Parkinsonism/7628819

### Summary of Updates

This version of the manuscript has been revised to substantially extend the experiments being conducted in the v1. The main updates are as follows: 1) adding illustrative Fig. 1 to better demonstrate the presented approach, 2) add strong baseline comparison, 3) showcase opportunity to solve the current limitations and possible bias in the SFM pretraining, 4) adding Chapter Implications. The changes and new experiments have addressed comments from the Editor with Information Processing and Management journal.

