## Supplementary Materials for "On Estimating Age and Gender from Parkinson’s Disease Diagnostic-Oriented Recordings Using Wav2Vec 2.0"

### Supplementary Material

#### Supplementary Methods - Bland–Altman analysis

Agreement between true chronological age and model-predicted age was assessed using Bland–Altman analysis. For each participant, the difference between predicted and true age was plotted against their mean. Mean bias (average prediction error) and 95% limits of agreement (mean bias ± 1.96 standard deviations) were computed for each speech task and cohort. This analysis complemented correlation-based metrics by quantifying systematic bias and the dispersion of individual prediction errors. No formal hypothesis testing was performed for Bland–Altman analysis, which was used descriptively to assess agreement.

#### Supplementary Figures

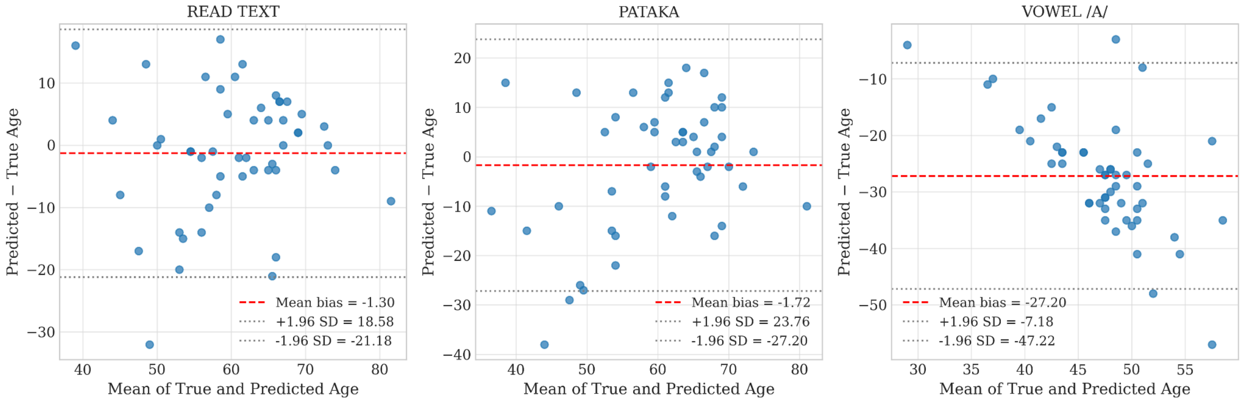


**Fig. S1.** Bland–Altman plots showing agreement between true chronological age and predicted age for the HC Dataset-1 cohort across three speech tasks.

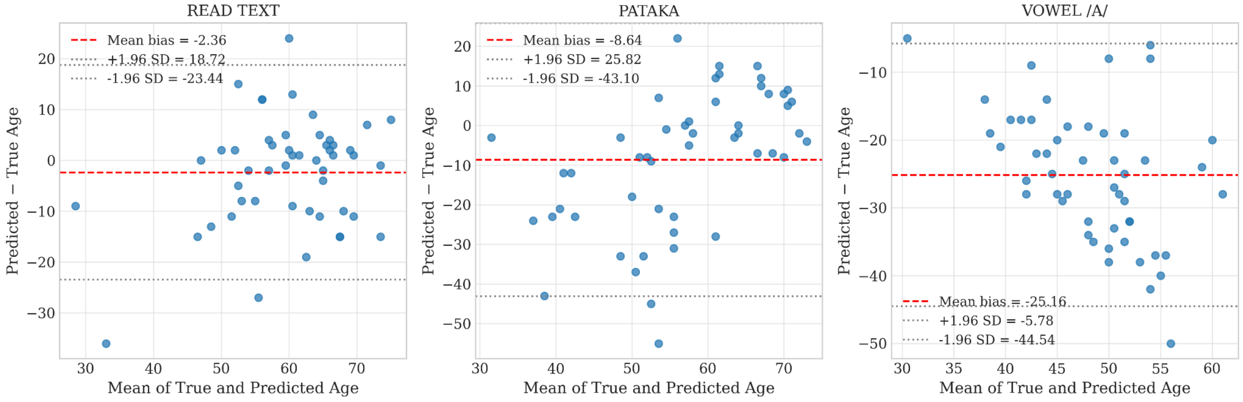


**Fig. S2.** Bland–Altman plots illustrating agreement between true and predicted age in PD Dataset-1 across three speech tasks.

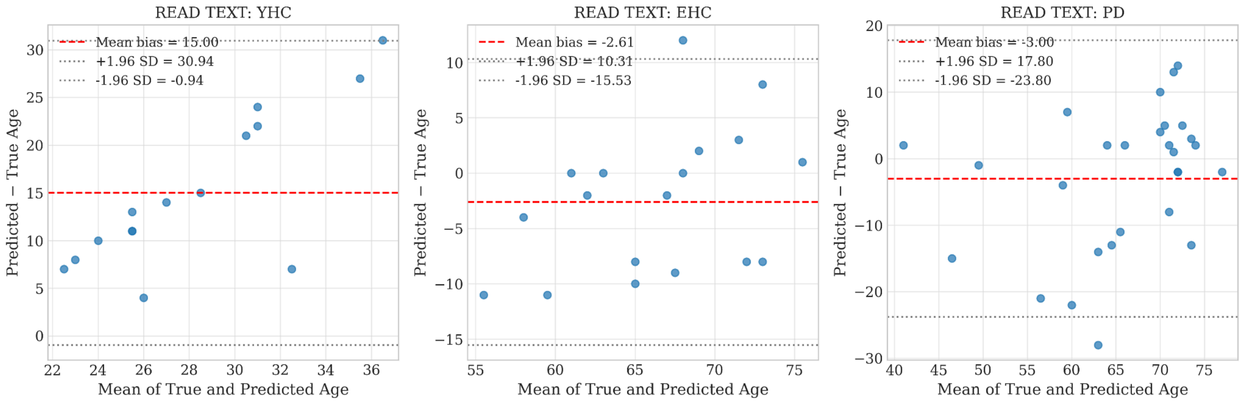


**Fig. S3.** Bland–Altman analysis of age prediction for the Italian dataset (Dataset-2), shown separately for YHC, EHC, and PD.
